# IL-5 blockade restores the bronchial epithelium and attenuates airway remodelling in severe asthma

**DOI:** 10.64898/2026.07.22.26358464

**Authors:** Onofrio Zanin, Alice J Eminton, Dasha Freydina, Varsha Kanabar-Raivadera, Isabella Drummond, Katie Horton, Juliette Phillips, Ravinder Dhillon, Jennifer Naftel, Wint Soe, Paddy Dennison, Laurie Lau, Jon Ward, Cornelia Blume, Emily J Swindle, Rocio T Martinez-Nunez, Hitasha Rupani

## Abstract

**Rationale:** Airway remodelling (AR) contributes to airflow limitation and poor symptom control in severe asthma. While anti-IL-5 therapy improves clinical outcomes in severe asthma, the cellular and molecular mechanisms underlying its effects on AR remain incompletely understood.

**Objectives.:** To determine whether IL-5 blockade directly modulates airway epithelial biology and contributes to attenuation of AR in severe asthma with eosinophilia (SAE).

**Methods.:** Patients with SAE underwent bronchoscopy before and after 24 weeks of anti-IL-5 therapy. Paired bronchial brushings (n=12) were analysed using single-cell RNA-sequencing. Histological features of AR were assessed in paired bronchial biopsies (n=16). Functional effects of IL-5 were investigated using wound healing assays in differentiated air-liquid interface (ALI) cultures.

**Measurements and Main Results.:** Anti-IL-5 treatment improved clinical outcomes without altering airway epithelial cellular composition. Differential gene expression was predominantly restricted to bronchial ciliated epithelial cells, which expressed *IL5RA*. ALI cultures showed IL-5Rα protein. Anti-IL-5 therapy induced a transcriptional signature in ciliated cells that opposed IL-5-responsive genes. Pseudotime analyses demonstrated preserved epithelial differentiation trajectories but altered programmes related to mucus regulation and ion transport. Cell-cell communication analyses revealed decreased T2-inflammatory processes alongside enrichment of epithelial repair and barrier integrity processes after treatment. Functionally, IL-5 directly impaired epithelial wound repair in ALI cultures. Histological assessment demonstrated increased epithelial E-cadherin expression and reduced sub-basement membrane thickness, extracellular matrix deposition and goblet cell hyperplasia in bronchial biopsies.

**Conclusions.:** IL-5 blockade modulates epithelial biology at transcriptional, functional and structural levels in SAE and is associated with improved epithelial integrity and reduced features of AR.

**Impact:** Our findings broaden current understanding of interleukin (IL)-5 biology by demonstrating that IL-5 blockade exerts direct effects on the bronchial epithelium in addition to its established effects on eosinophilic inflammation in asthma. Through single-cell analyses of human bronchial samples, complemented by functional and histological validation we provide novel mechanistic insight into how IL-5 blockade influences epithelial repair, barrier integrity and airway remodelling. These findings advance understanding of severe asthma pathogenesis, provide evidence that structural disease is modifiable and may inform future approaches to disease modification.

**At a Glance Commentary:** *Scientific knowledge on the subject:* Interleukin-5 (IL-5) is a key driver of eosinophilic inflammation in severe asthma and anti-IL-5 therapies improve clinical outcomes. Emerging evidence suggests that IL-5 blockade may also improve features of airway remodelling. However, the cellular mechanisms underlying airway structural improvements and the effects of IL-5 blockade on airway structural cells remain poorly understood.

*What this study adds to the field:* Using paired bronchial samples obtained before and after anti-IL5 treatment, we demonstrate that IL-5 blockade directly modulates bronchial epithelial cell biology. We identify ciliated epithelial cells as an IL-5 responsive population and show that anti-IL-5 therapy alters transcriptional programmes in airway epithelium associated with repair, barrier integrity and airway remodelling. IL-5 blockade shifts cell-cell communication away from inflammation pathways towards pathways supporting epithelial repair. Functionally, IL-5 directly impairs wound healing of differentiated airway epithelial cells *in vitro* and IL-5 blockade enhances epithelial integrity and reduces remodelling-associated structural changes *in vivo*. Our findings broaden current understanding of IL-5 biology by demonstrating that the effects of IL-5 blockade extend beyond eosinophil suppression to include direct effects on biological pathways in epithelial cells involved in repair, barrier integrity and airway remodelling in severe asthma.

## Introduction

Asthma is a common chronic inflammatory airway disease. Central to asthma pathogenesis is the bronchial epithelium, which forms the primary interface between the host and the external environment, integrating barrier protection with regulation of immune homeostasis^1^. It comprises a complex network of specialised cell types including stem cell-like basal cells, ciliated cells and goblet cells. The importance of the epithelium in asthma is highlighted by genome-wide association studies identifying numerous asthma susceptibility genes expressed within bronchial epithelial cells^2,3^.

The bronchial epithelium initiates and amplifies type-2 (T2) inflammation, the dominant inflammatory endotype in most patients with severe asthma^4^. Following exposure to environmental challenges, epithelial damage triggers the release of alarmins that activate T-helper (Th2) cells, mast cells and group 2 innate lymphoid cells (ILC2s) ^1,5^. These cells secrete T2 cytokines including interleukin (IL)-5, which drives the recruitment, activation and survival of eosinophils within the airway, alongside IL-13 and IL-4, which promote mucus hypersecretion, allergen-specific IgE production and airway hyperresponsiveness^1,5^. Eosinophils are considered key effector cells in severe asthma and elevated eosinophil levels in blood and airway compartments are associated with disease exacerbations and lung function decline.

In asthma, epithelial barrier disruption and dysfunction initiates and amplifies inflammatory signalling and perpetuates tissue injury, driving airway remodelling (AR), an aberrant and exaggerated repair response to chronic inflammation and repeated bronchoconstriction^6^. AR is characterised by mucosal oedema, mucus hypersecretion, airway smooth muscle hypertrophy/hyperplasia and subepithelial extracellular matrix (ECM) deposition, leading to progressive airflow limitation and poor symptom control. While airway inflammation is generally responsive to corticosteroids, AR has been considered largely irreversible, highlighting a major unmet need in the management of severe asthma.

Therapeutic targeting of IL-5 with biologics such has mepolizumab reduces exacerbations and improves lung function in patients with severe asthma^7^. Emerging evidence indicates that mepolizumab may also modulate AR with reported improvements in epithelial integrity, reductions in basement membrane (BM) thickness^8,9^ and decreased deposition of ECM proteins^10^. However, the mechanisms underlying these effects remain incompletely defined, particularly regarding the impact of IL-5 blockade on structural cells expressing the IL-5 receptor, such as ciliated epithelial cells^11^.

We hypothesised that IL-5 blockade modulates AR by direct reprogramming of the bronchial epithelium. We tested this hypothesis by applying single-cell, cellular and immunohistochemical analyses of paired pre- and post-anti-IL-5 treatment human bronchial brushings and biopsies. Our findings provide novel mechanistic insight into bronchial epithelial responses to IL-5 blockade and how anti-IL-5 therapy may modulate structural disease processes beyond eosinophil suppression.

## Methods

Full details are provided in the Supplementary Material and Methods.

### Patient population

Sixteen non-smoking patients with severe asthma with eosinophilia (SAE, ≥300 cells/μL within the last 12 months), a history of ≥3 exacerbations in the previous year despite high dose inhaled corticosteroid and long-acting beta-2 agonist (ICS/LABA) therapy and due to start mepolizumab therapy (100 mg subcutaneously every 4 weeks; GSK, Barnard Castle, County Durham, England) as part of their standard clinical care were recruited. Participants underwent clinical characterisation and bronchoscopy with airway sampling before and after 24 weeks of mepolizumab (anti-IL-5 therapy herein) treatment. The study was approved by Cambridge East Research Ethics Committee: REC reference 21/EE/0228, IRAS 304497. All participants provided written informed consent.

### Flexible bronchoscopy and sample preservation

Participants underwent bronchoscopy conducted according to British Thoracic Society guidelines^12^. Bronchial brushes and biopsies were collected, processed and cryopreserved or fixed respectively.

### Single cell processing

Bronchial brushes were thawed and immediately tested for viability, presence of clusters and debris and diluted following 10X Genomics instructions.

### Western blotting

Protein samples were reduced, denatured, resolved by electrophoresis, blotted on nitrocellulose membranes, blocked and incubated with antibodies.

### Pathway analysis

We compared public resources using Harmonizome 3.0 (maayanlab.cloud/Harmonizome)^13^.

### Bronchial epithelial cell (BEC) culture

*Generation of BEAS-2B-IL5RA:* BEAS-2B cells stably expressing IL-5Rα were generated by lentiviral transduction of the *IL5RA* cDNA (Origene). *BCi scratch-wound assay*. BCi-NS1.1 (BCi herein) cells were cultured at the air liquid interface (ALI) for 28 days +/-IL-5 (0.5 ng/mL). Scratch wound repair was visualised over 24h using light microscopy.

### Histology and Immunohistochemistry

Endobronchial biopsy sections were fixed in 4% paraformaldehyde, paraffin embedded, processed and stained as previously described^14^.

### Code

All code is available at https://github.com/martinez-nunez-lab/iris_sccb

### Statistical Analysis

Clinical parameters, immunohistochemistry and scratch wound assays statistical analyses were performed in GraphPad Prism v10.3.1 (GraphPad Software Inc, San Diego, CA). *P* ≤ 0.05 marked statistical significance.

## Results

### Clinical improvement after anti-IL-5 therapy

At baseline, participants (9 male, 7 female; median age [IQR] 64 [54,68] years) had a high burden of disease (annualised exacerbate rate 5 [4,7], asthma control questionnaire 6 [ACQ6]: 2.65 [2.03,3.28]) and impaired lung function (FEV_1_ % predicted 63.50% [58.75,83.00]). All received high-dose ICS (>1000mcg beclometasone dipropionate equivalent) plus additional controller therapy. Mepolizumab significantly improved clinical outcomes and reduced blood and bronchoalveolar lavage eosinophils (see Table S1 in the Online Supplement).

#### IL-5 blockade does not change the cellular composition of the bronchial epithelium

Paired bronchial brushings from 12 participants were processed for single cell RNA sequencing (Figure 1A). After filtering and batch effect correction, 77,033 cells were included in the analysis, similarly distributed pre- and post-treatment (38,972 and 38,061 cells respectively, see Figure S1A in the Online Supplement). Overall, 38,606 genes and 11 cell populations were identified including epithelial and immune cell types (Figure 1B-1C). Within BEC populations we identified basal, club, ciliated, deuterosomal, goblet and mucociliated cells and ionocytes, with ciliated cells being the most abundant. Amongst immune cells, T cells, macrophages, dendritic cells and mast cells were identified. Eosinophils did not withstand sample processing and are thus absent from our analysis.

**Figure 1.**
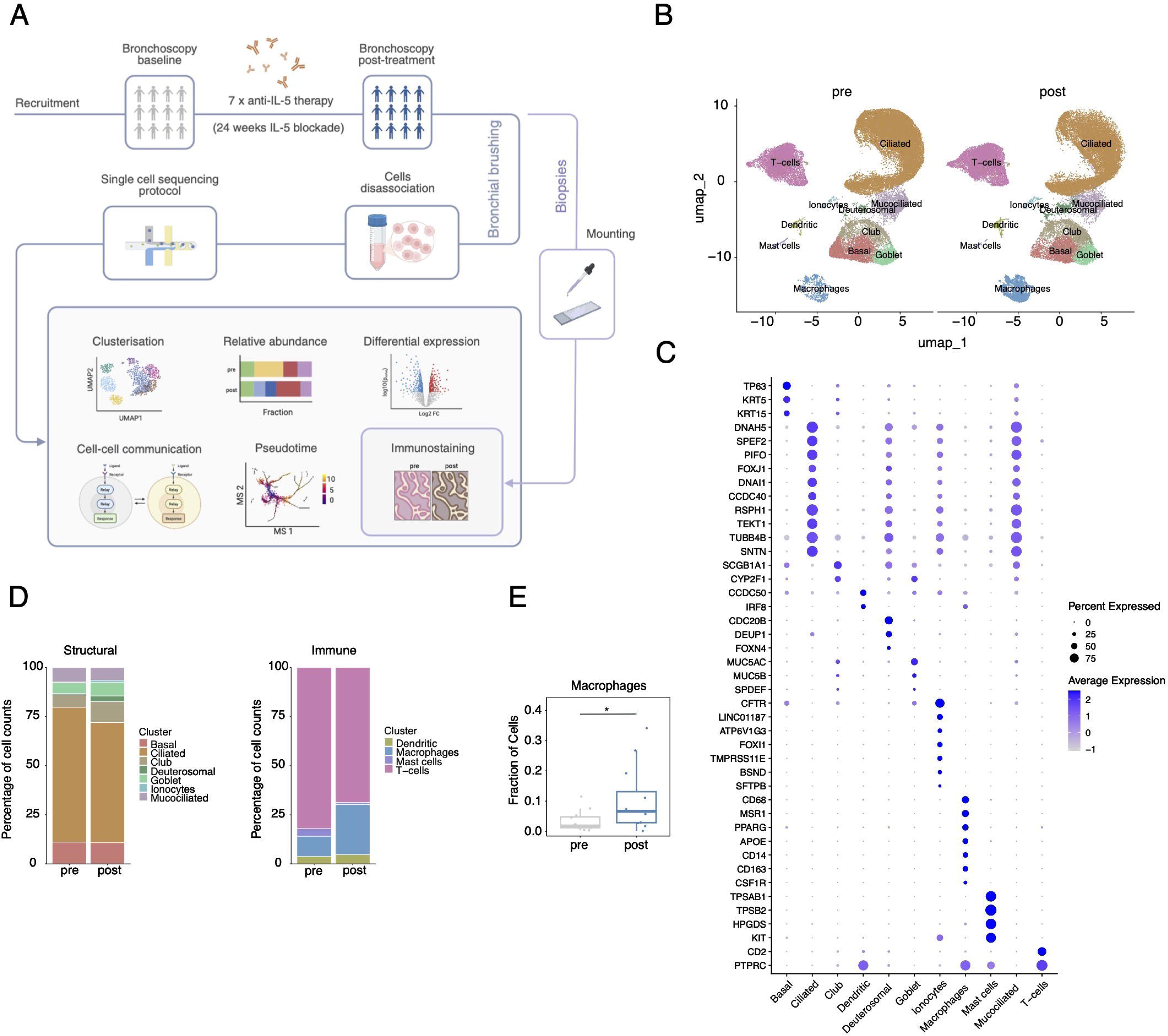
**A.** Schematic depicting the experimental approach including sampling, processing and analysis. 16 patients underwent bronchoscopy and airway sampling pre- and after 24 weeks of anti-IL-5. Samples where cryopreserved, processed for scRNA-sequencing and analysed to investigate cellular composition (clusterisation/relative abundance), differential gene expression per cell type, cell to cell communication and pseudotime variations. Bronchial biopsies were immunostained. **B.** UMAP plots showing the cellular composition of the bronchial brushings pre- and post-treatment. **C.** Dot-plot listing the marker genes employed for cluster characterisation and annotation. **D**. Stacked bar graph showing the cellular proportions of epithelial and immune cell types found in the bronchial brushings pre-and post-treatment. **E.** Box plot showing an increase in the bronchial brushing macrophage fraction post-treatment; * = p<0.05.

Anti-IL-5 treatment did not change the proportion of most cellular populations isolated (Figure 1D, see Figure S1B in the Online Supplement) except macrophages which were increased post-treatment and enriched in M2-skewed activation (*CD163*, *MRC1* (CD206), *CLEC10A* (CD301)) (Figure 1E, see Figure S1C in the Online Supplement). Mast cells showed a trend towards decreased proportions post-treatment (see Figure S1B in the Online Supplement). Overall, our results suggest that the clinical improvements observed following anti-IL-5 therapy are not accompanied by major shifts in bronchial epithelial composition.

#### Anti-IL-5 treatment distinctively alters the transcriptional profile of ciliated epithelial cells

Studies have demonstrated that anti-IL-5 therapy induces changes in nasal epithelial samples by bulk transcriptomics^7,15,16^ where mixed cellular populations are present. To define cell-specific transcriptional responses to IL-5 blockade in BECs, we compared each identified cell population pre- and post-treatment using pseudobulk analysis.

Differentially expressed genes (DEGs, p-adj<0.05) were observed exclusively within BEC populations, specifically ciliated cells (325 genes), ionocytes (23 genes) and basal cells (15 genes) (Figure 2A). Twenty-nine percent of DEGs in ciliated cells were upregulated while 71% were downregulated. Twenty DEGs identified in ciliated cells co-occurred with the biological term ‘remodelling’^13^ (Figure 2B, left and see Tables S2-4 in the Online Supplement), including downregulation of *SMAD2* expression, a transcription factor that promotes AR and BM thickening in asthma^17^. Overall, ∼11% of all DEGs in ciliated cells were predicted to be SMAD2-dependent (Figure 2B, right). Several DEGs were associated with cell-cell junction pathways (see Figure S2A in the Online Supplement), suggesting that anti-IL-5 therapy influences both structural remodelling and barrier integrity.

**Figure 2.**
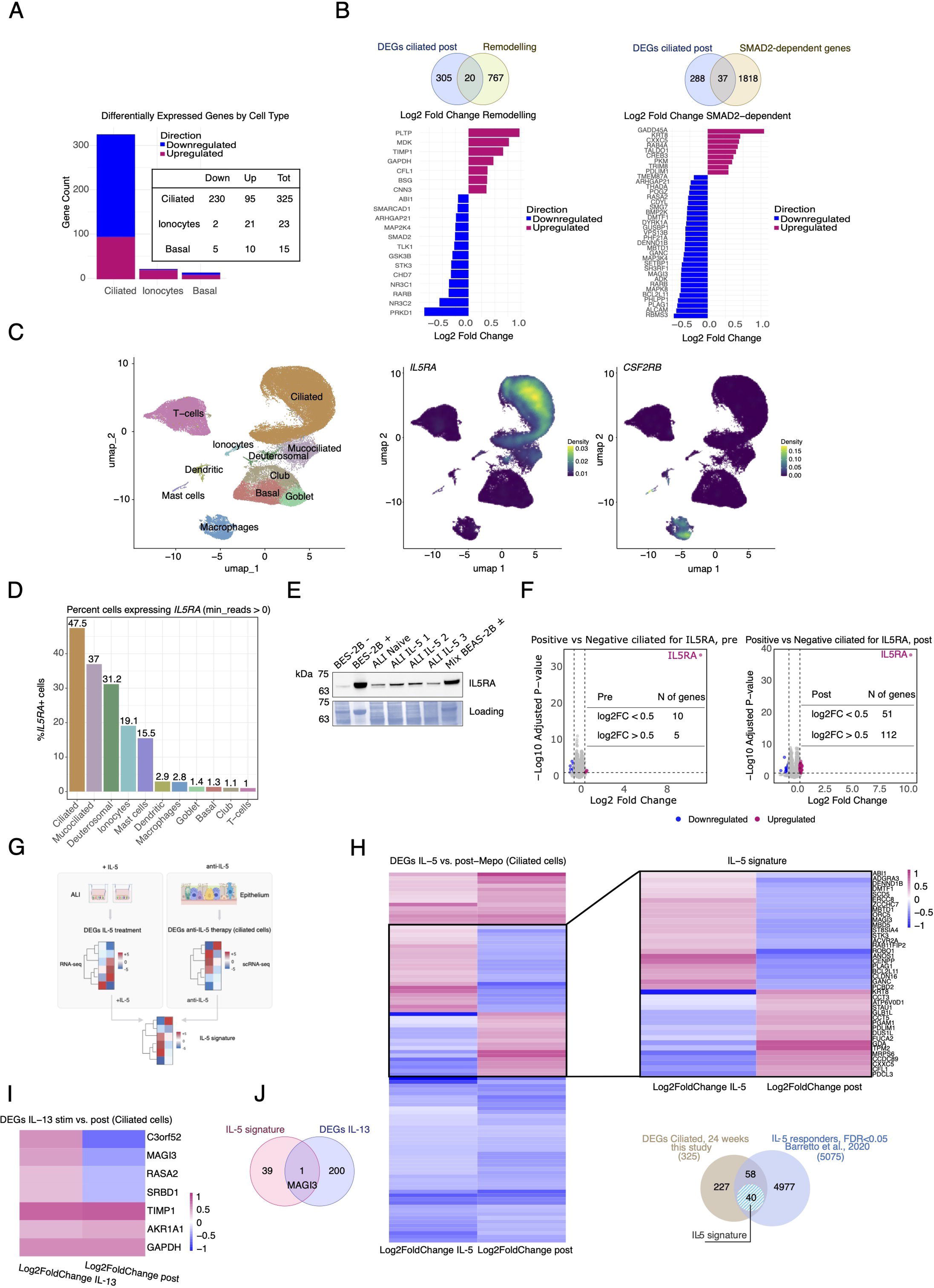
**A.** Stacked bar plot showing the number of statistically significant differentially expressed genes (DEGs) (adjusted p-value (padj) < 0.05). Internal table shows the number of DEGs downregulated (Down), upregulated (Up) and total (Tot). **B**. Venn diagrams showing the overlap between DEGs observed in ciliated cells and genes within the biological term “remodelling” (left) and “SMAD-2-dependent” (right). Bar plots listing specific genes in the biological term “remodelling” (left) and “SMAD-2-dependent” (right)-genes are displayed on the y-axis and ranked according to the magnitude of differential expression (log2 Fold change) on the x-axis; blue: downregulated; red, upregulated. **C**. Left: UMAP plot showing the annotated clusters and cell types. Middle: UMAP highlighting the cells expressing *IL5RA.* Right: UMAP highlighting the cells expressing *CSF2RB.* **D.** Bar plot showing the percentage of cells per cell type expressing *IL5RA*. **E**. Western blot for IL-5Rα expression in wild type BEAS-2B cells (BEAS-2B -, lane 1), BEAS-2B cells overexpressing *IL5RA* (BEAS-2B +, lane 2), unstimulated primary ALI cultures (ALI naïve, lane 3) and IL-5 treated ALI cultures (ALI IL-5 1, 2, 3. Lanes 4, 5, 6) and a mix of wild type BEAS-2B and transgenic BEAS-2B cells (Mix BEAS-2B ±, lane 7). Ponceau staining shown as a loading control. **F.** Volcano plots showing DEGs between *IL5RA*-positive and *IL5RA*-negative ciliated epithelial cells pre-treatment (2941 DEGs) and post-treatment (3801 DEGs). DEGs with -0.5 > log_2_fold change > 0.5 are highlighted (blue downregulated and red upregulated). **G.** Schematic of the comparison of DEGs of ALI cultures exposed to IL-5^11^ and DEGs in the ciliated cells post IL-5 blockade. **H**. Left: heatmap comparing IL-5 responsive genes identified in ALI cultures^11^ with DEGs in ciliated cells from brushings following IL-5 blockade. Genes exhibiting reciprocal regulation following IL-5 blockade are highlighted and termed the ‘IL-5 signature’. Bottom right: Venn diagram showing the overlap between DEGs identified following IL-5 stimulation of ALI cultures^11^, DEGs identified in ciliated epithelial cells following IL-5 blockade *in vivo*, and the epithelial IL-5 signature. **I.** Heatmap showing the comparison of DEGs in ciliated cells obtained from exposure of ALI cultures to IL-13^20^ and DEGs in ciliated cells from brushings post anti-IL-5 blockade. **J.** Venn diagram showing the overlap between DEGs induced by IL-5 treatment^11^, IL-13 treatment^20^ and the IL-5 signature genes.

Anti-IL-5 treatment may exert both direct effects through IL-5 receptor signalling, and indirect effects via eosinophil reduction. Therefore, we investigated the expression of the IL-5 receptor subunits, *IL5RA* and *CSF2RB*, across all cell populations. Expression of both subunits was heterogenous (Figure 2C) with *IL5RA* mRNA predominantly detected in ciliated cells (47.5%), with lower expression in ionocytes (19%), basal cells (1.3%), mast cells (15.5%) and T cells (1%) (Figure 2D). *CSF2RB* expression was undetectable in BECs and restricted to immune cells, including mast cells, dendritic cells and macrophages, consistent with its role as the shared β-chain of GM-CSF^18–20^ (Figure 2C).

We confirmed IL-5Rα protein expression in primary BECs differentiated at the ALI, which contain ciliated cells (Figure 2E). As controls, BEAS-2B cells constitutively transduced with *IL5RA* showed a strong protein band at the expected molecular weight, whereas wild-type BEAS-2B cells showed no detectable specific signal (Figure 2E). Similarly, IL-5Rα protein was detected in both BCi cell monolayers and differentiated ALI cultures but was absent in fibroblasts from patients with severe asthma (see Figure S2B in the Online Supplement). Consistent with these findings, bulk RNA-sequencing of fibroblasts outgrown from bronchial biopsies pre- and post-treatment showed no expression of *IL5RA* and no significant transcriptional response to anti-IL-5 therapy (see Figure S2C in the Online Supplement).

To determine whether *IL5RA* expression influenced transcriptional responses to anti-IL-5 treatment, we compared *IL5RA*^+^ and *IL5RA*^-^ ciliated cells before and after treatment. Pre-treatment, 15 DEGs were identified between these cell populations, whereas post-anti-IL-5 treatment this number increased to 163 DEGs (|log2 fold change| >0.5, p-adj<0.05, Figure 2F and see Figure S2D in the Online Supplement). These findings suggest that *IL5RA* presence is associated with transcriptional responses to IL-5 blockade in ciliated epithelial cells.

To further assess whether anti-IL-5 therapy directly modulates BECs, we compared post-treatment DEGs with a published IL-5-induced gene expression dataset from ALI cultures^11^ (Figure 2G). As a comparator, we performed the same analysis using DEGs from ciliated cells upon IL-13 stimulation of ALIs^21^, given the established effects of IL-13 on gene expression in the epithelium in asthma.

This analysis identified a subset of DEGs in ciliated cells whose expression changed in the opposite direction post-anti-IL-5 therapy compared with IL-5 stimulation *in vitro*, indicating reversal of IL-5-dependent changes (Figure 2H). This inverse trend was observed in approximately 40% of genes that were both IL-5 responsive and differentially expressed following treatment, which we termed the ‘epithelial IL-5 signature’. Among these were *CFL1*-restoring this actin-binding protein improves barrier integrity^22^. In contrast, comparison with the IL-13-dependent signature^21^ identified 7 overlapping genes, of which a single gene (*MAGI3*, Figure 2I) was shared with the epithelial IL-5 signature. Although fewer DEGs were identified in basal cells and ionocytes following treatment, both populations also exhibited distinct IL-5-associated transcriptional signatures (see Figures S2E and S2F in the Online Supplement).

Together these findings indicate that transcriptional changes in BECs upon anti-IL5 therapy are highly specific to IL-5-dependent pathways, identify ciliated epithelial cells as a novel IL-5 responsive population and demonstrate that anti-IL-5 therapy directly modulates bronchial epithelial biology.

#### Anti-IL-5 therapy modulates epithelial activation states without altering differentiation trajectories

We next investigated whether IL-5 blockade altered epithelial differentiation state or lineage trajectories using pseudotime analysis.

Analysing the pseudotime of key epithelial markers enabled determining the progression and fate of specific epithelial populations (represented in Figure 3A). Basal cells showed the lowest pseudotime (high *TP63* expression, root of trajectory), consistent with a stem cell/progenitor phenotype (Figures 3B-3C), while ionocytes occupied a distinct side branch and displayed the highest pseudotime, representing a specialised trajectory. All other epithelial cell types displayed intermediate pseudotime (Figures 3B-C). The behaviour of these lineages was confirmed using specific marker genes for basal cells and ionocytes (Figure 3D). Overall, IL-5 blockade did not significantly alter epithelial differentiation trajectories (Figure 3B) or cell fate decisions (Figure 3C) in BECs.

**Figure 3.**
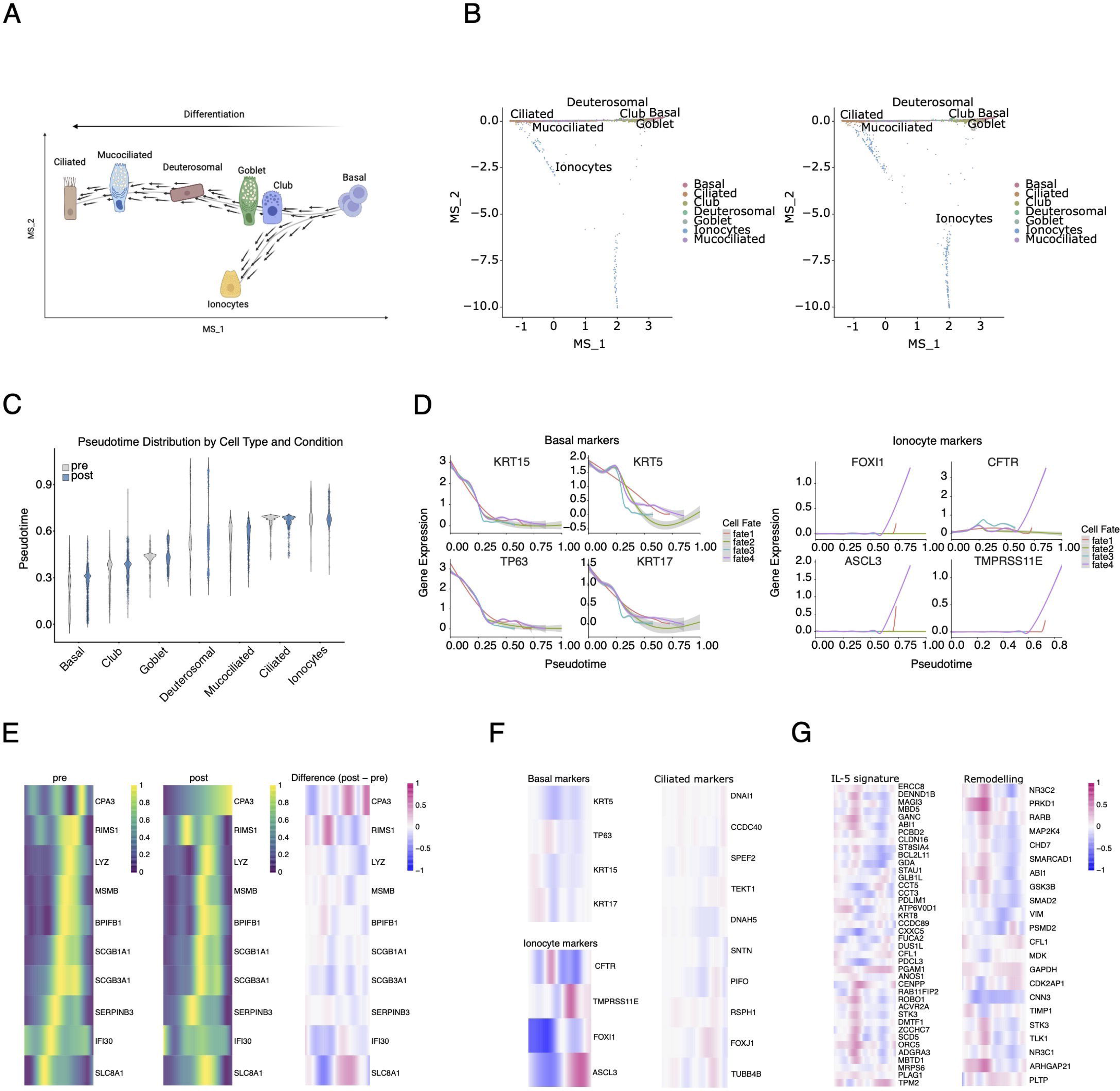
**A**. Schematic depicting the principle of the cell fate path on the diffusion map shown in B. **B.** Diffusion map from dimensionality reduction of trajectory inference of the epithelial cell fraction of bronchial brushings analysed for scRNA-seq pre (left) and post (right) IL-5 blockade. **C.** Violin plot showing the pseudotime calculated per cell type between pre and post treatment. **D.** Gene trend curves showing the gene expression of gene markers along the pseudotime for basal cells and ionocytes. **E.** Heatmaps showing the expression levels of selected genes along the pseudotime pre- and post-treatment and the measured difference post treatment for the top 10 variable genes (red upregulated post treatment or downregulated pre-treatment, blue downregulated post-treatment or upregulated pre-treatment). **F.** Heatmaps showing the measured difference along the pseudotime post-treatment for ionocytes, basal cell and ciliated cells markers, the genes listed in the IL-5 signature and the genes occurring with the biological term “remodelling” (red upregulated post treatment or downregulated pre-treatment, blue downregulated post treatment or upregulated pre-treatment).

In contrast, anti-IL-5 therapy selectively altered the expression along the pseudotime of genes associated with epithelial function and inflammation. Among the most variable genes in epithelial cells, *CPA3* and *SLC8A1*, shifted towards later pseudotime following treatment, whereas *RIMS1* shifted earlier (Figure 3E). *SLC8A1* and *RIMS1* encode proteins implicated in mucus regulation^23,24^, while epithelial *CPA3* expression is associated with IL-4/IL-13 driven JAK/STAT6 signalling in asthma^25^, suggesting modulation of inflammatory and mucus-associated epithelial programmes following anti-IL-5 therapy.

Anti-IL-5 treatment also modified the expression along the pseudotime of key epithelial markers. In ionocytes, *CFTR*, which regulates ion flux required for adequate mucus hydration and ciliary function^26^, shifted towards earlier pseudotime states. *TMPRSS11E* shifted towards later pseudotime while ionocyte transcription factors *FOXI1* and *ASCL3*^27^ were restricted to intermediate and later pseudotimes (Figure 3F). By comparison, the pseudotime distribution of basal and ciliated cell markers remained unchanged following treatment (Figure 3F).

Finally, remodelling-associated genes (Figure 2B) and the epithelial IL-5 signature (Figure 2I) demonstrated a modest shift towards earlier pseudotime states following anti-IL-5 therapy (Figure 3G), consistent with restoration of epithelial homeostasis through altered activation states rather than changes in lineage composition.

Collectively, these findings indicate that anti-IL-5 therapy does not alter epithelial differentiation trajectories in severe asthma, but instead modulates specific transcriptional processes associated with epithelial activation, mucus regulation, ion transport and AR.

#### IL-5 blockade improves epithelial communication networks associated with airway remodelling and repair

In the airways, BECs exist within a complex network of interactions rather than as isolated cell populations. To investigate whether anti-IL-5 therapy influences intercellular signalling, we analysed cell-to-cell communication patterns using CellChat^28,29^.

We found extensive cell-cell networking before and after anti-IL-5 treatment (see Figure S3A in the Online Supplement) indicating active communication between epithelial and immune cell populations with an overall increase in the number and strength of interactions following anti-IL-5 therapy (Figure 4A). Among epithelial cell populations exhibiting DEGs following treatment, anti-IL-5 treatment increased both the number and strength of interactions between ciliated cells and basal cells and between ciliated cells and ionocytes, while communication within basal cells was reduced (Figure 4B).

**Figure 4.**
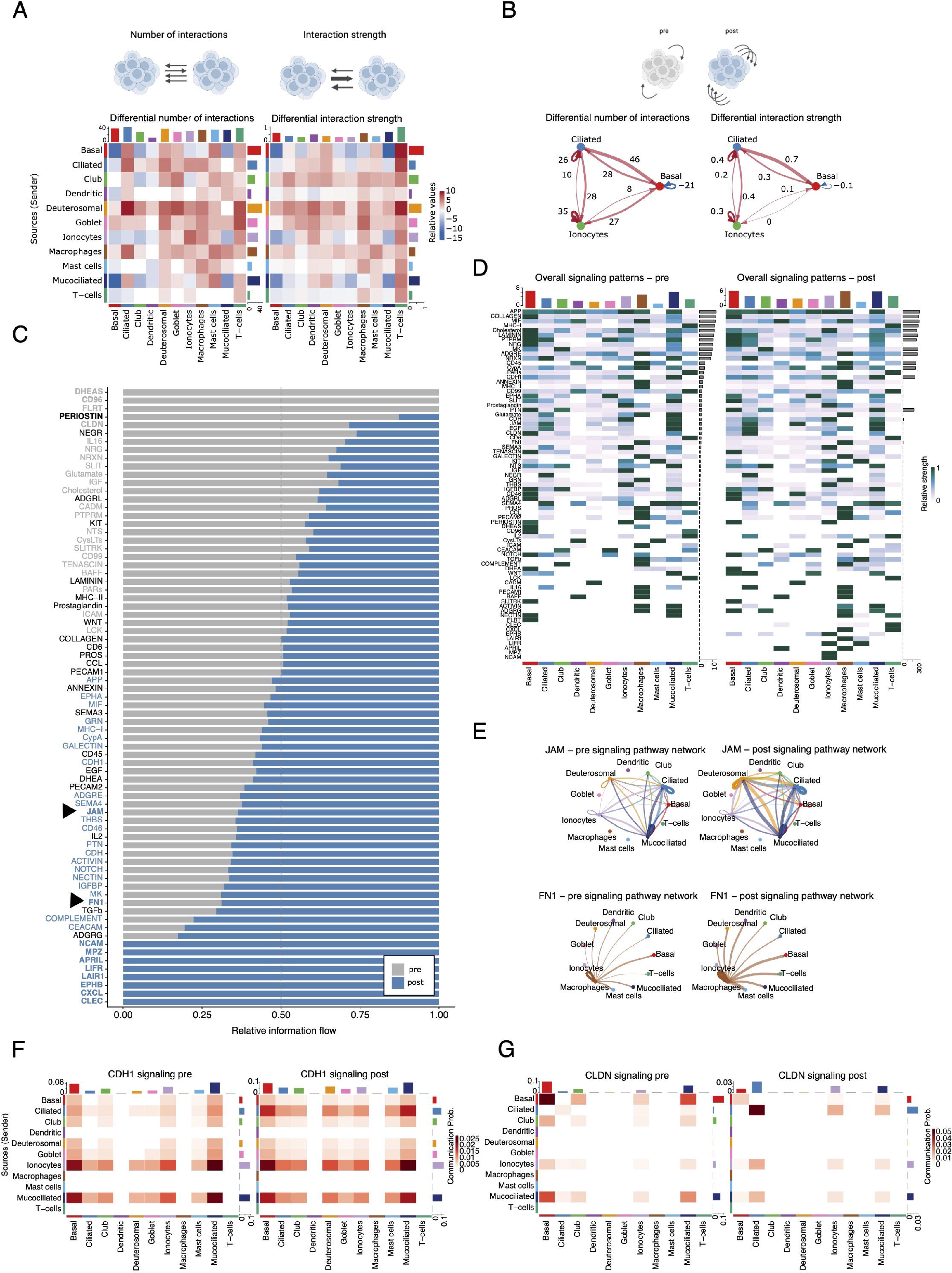
**A.** Schematic illustrating the principle of interaction number and interaction strength. Heatmaps show the differential number of interactions (left) and differential interaction strength (right) comparing pre- and post-treatment. Red indicates increased interactions post-treatment and blue indicates decreased interactions. **B.** Schematic illustrating the concept of differential number and strength of interactions pre and post treatment. Circle plots showing the differential number of interactions and the differential interaction strength between ciliated, basal and ionocytes pre-and post-treatment. **C.** Stacked bar plot showing cell-cell communication pathways and information flow between pre and post treatment. Relevant pathways are highlighted or indicated by black triangles. **D.** Heatmap showing overall signal intensities of all cell-cell signalling pre- and post-treatment. **E.** Circle plot relative to communication networking for Junctional Adhesion Molecule (JAM, top) and fibronectin (FN1, bottom) pathways pre- and post-IL-5 blockade. **F-G.** Heatmaps showing the communication probability pre- and post-treatment per cell type related to E-cadherin (CDH1, **F**) and claudin (CLDN, **G**) pathways.

To explore the biological relevance of these altered interactions, we mapped intercellular communication patterns to signalling pathways (Figure 4C). Consistent with attenuation of T2 inflammation following IL-5 blockade, periostin (POSTN), a marker of T2 inflammation in asthma^30^, demonstrated reduced pathway activity after treatment. Pathways selectively enriched pre-treatment were related to inflammation (DHEAS, CD96) ^31–33^ and cell adhesion and fibroblast growth factor signalling (FLRT) ^34^. Cell-cell communication pathways selectively enriched following anti-IL-5 treatment were predominantly associated with epithelial repair, barrier integrity and immune modulation. These included NCAM, a marker of neuroendocrine cells which are rare but pivotal in airway epithelium^35^, MPZ, APRIL^36^, LIFR, LAIR1, EPHB^37^, CXCL and CLEC^38^.

AR-associated pathways, including collagen and laminin signalling, were prominent (Figures 4C-4D) and involved intense communication across multiple cell populations (see Figure S3 in the Online Supplement). Collagen cell-cell communication was predominantly mediated through COL4A5-dependent interactions involving CD44 and integrin receptors (see Figure S3 in the Online Supplement), highlighting active epithelial-matrix communication networks. Following anti-IL-5 therapy, pathways linked to epithelial integrity and homeostasis were enhanced, including junctional adhesion molecule (JAM), fibronectin (FN1) (Figure 4E), cadherin (CDH1) and claudin (CLDN) signalling networks, particularly within ciliated cells and ionocytes (Figures 4F-4G). Collectively, these finding indicate IL-5 blockade induced broad reorganisation and enhancement of cell-cell communication and shifted intercellular signalling away from inflammation-associated pathways towards pathways supporting epithelial repair, barrier integrity and tissue homeostasis.

#### IL-5 directly impairs epithelial wound repair and IL-5 blockade attenuates airway remodelling

Next, we investigated whether IL-5 directly influences epithelial repair capacity. BCi cells were differentiated in the absence or presence of IL-5 (0.5 or 10 ng/mL), modelling the microenvironment of SAE, before undergoing scratch wound assays (Figure 5A). ALI cultures exposed to 10 ng/mL IL-5 during differentiation and repair exhibited significantly impaired wound repair at 12, 18 and 24 hours compared with untreated controls and cells exposed to 0.5 ng/mL IL-5 (Figure 5B, left). This effect was abolished when IL-5 was removed immediately prior to wounding (Figure 5B, right), mimicking therapeutic IL-5 blockade *in vitro*. These findings provide functional evidence that IL-5 directly impairs epithelial repair responses and supports our previous transcriptomic and cell-cell communication analyses indicating restoration of epithelial repair pathways post-IL-5 therapy.

**Figure 5.**
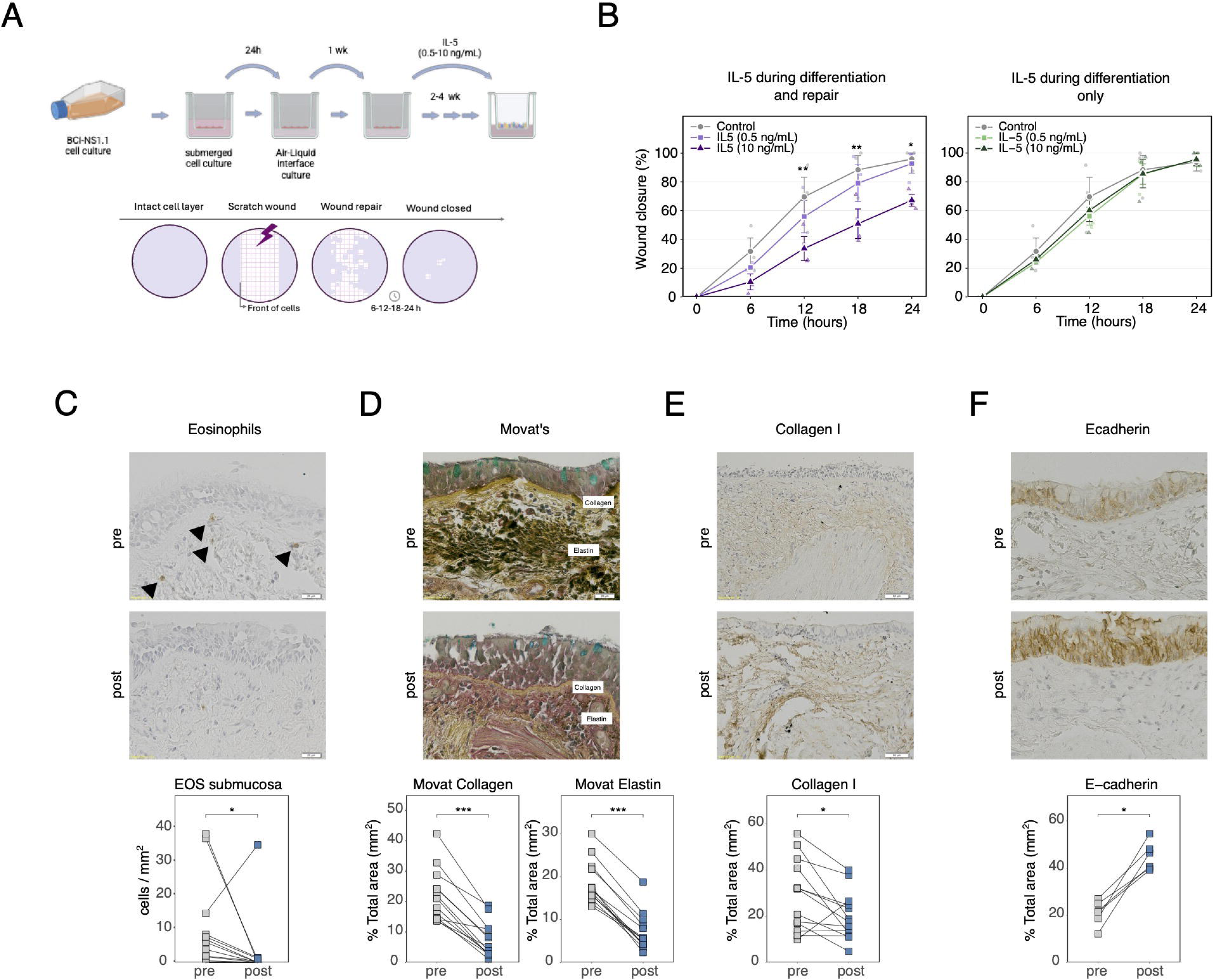
**A.** Schematic illustrating the *in vitro* scratch wound healing experimental design. **B.** Differentiated BCi cells exposed to IL-5 (0.5-10 ng/ml) during differentiation and repair (left hand side) or differentiation only (right hand side). Results are mean ± s.d., n=3 (in triplicate). **C-F.** Top images: representative bronchial biopsy slices pre- and post-IL-5 blockade showing eosinophils (C, black arrows), Movat’s pentachrome staining for total collagen (yellow staining), elastin(black staining) and mucin (blue staining) (D), Collagen I (E) and E-cadherin (F). Bottom: Dot plots showing the quantification of IHC/Movat’s pentachrome staining in C-F.

To determine if the molecular and cellular changes following anti-IL-5 therapy translated into structural tissue alterations *in vivo*, we analysed histological markers of AR and epithelial integrity in paired bronchial biopsies. As expected, anti-IL-5 therapy significantly reduced tissue eosinophilia (Figure 5C). Anti-IL-5 therapy was associated with significant improvement in histological features of AR with decreased ECM deposition of collagen and elastin (Figure 5D) and collagen I (Figure 5E), and reduced goblet cell hyperplasia and sub-BM thickness (Figure 5D and see Figure S4 in the Online Supplement). In parallel, expression of E-cadherin, a component of adherens junctions in the epithelial barrier was significantly increased (Figure 5F) indicating improved epithelial barrier integrity.

Collectively, these findings demonstrate that IL-5 blockade is associated with structural improvement of the airway epithelium and attenuation of AR-related changes in severe asthma, paralleling restoration of epithelial molecular signatures and repair responses.

## Discussion

Here we demonstrate that IL-5 blockade directly modulates BEC biology in severe asthma. By integrating single-cell transcriptomics, functional epithelial assays and histological analyses, we show that IL-5 blockade is associated with specific gene expression changes in the bronchial epithelium and attenuation of AR in severe asthma. These findings broaden the current understanding of IL-5 biology and identify the bronchial epithelium as an important target of anti-IL-5 therapy.

Our data provides evidence for direct IL-5 signalling in BECs *in vivo*, consistent with previous work^11^. Detection of *IL5RA* at mRNA and protein levels, together with identification of IL-5-responsive genes whose expression was selectively reversed following anti-IL-5 treatment supports epithelial responsiveness to IL-5. Importantly, there was limited overlap between the epithelial IL-5 signature and IL-13-dependent genes, suggesting that the detected changes do not solely reflect global suppression of T2-inflammation, but rather modulation of specific IL-5-dependent genes in BECs. Similar epithelial reprogramming has been reported following anti-IL-5 therapy in chronic rhinosinusitis with nasal polyps^39^, supporting the concept that structural airway cells are direct targets of IL-5 blockade. Further support comes from pulmonary fibrosis where increased epithelial *IL5RA* expression has been associated with reduction in E-cadherin expression, upregulation of remodelling-associated genes and ECM deposition, whereas IL5RA inhibition attenuated these changes^40^. These findings are consistent with our observations that IL-5 impairs epithelial repair, whereas IL-5 blockade promotes epithelial integrity and attenuates AR.

We found no detectable *CSF2RB* in ciliated cells and ALI cultures. The inhibitory effect of IL-5 on epithelial wound repair together with the IL-5 signature suggest that IL-5 signalling in BECs likely differs from classical IL-5 receptor signalling in eosinophils and other immune cells^20^. Sohail et al reported expression of *IL5RA* in nasal epithelial cells in the absence of *CSF2RB*^41^, although our data sugggest that IL-5 directly signals in BECs. Future studies should focus on elucidating how IL-5 signals within epithelial cells.

The predominance of downregulated transcripts following treatment mirrors findings in the nasal epithelium of patients receiving mepolizumab^42^ suggesting global gene expression repression, possibly via epigenetic mechanisms. The reduction in *SMAD2* is particularly relevant given the central role of TGF-β/SMAD signalling in ECM deposition and fibrosis^17^, and suggests that SMAD2 may contribute to the transcriptional programme driven by IL-5 blockade to attenuate profibrotic processes in AR.

Further novel mechanistic insight on how IL-5 blockade may influence AR comes from cell-cell communication analyses, which demonstrated a shift away from inflammatory pathways (e.g. POSTN, DHEAS, CD96) ^30–33^ towards pathways associated with epithelial repair, barrier integrity and tissue homeostasis. Consistent with this, a previous study has reported increased expression of junctional genes in nasal epithelium following mepolizumab treatment^42^. Additionally, pseudotime of genes related to mucus and ion transport shifted post-treatment. Together, these observations suggest that structural improvements following anti-IL-5 therapy are not solely related to suppression of eosinophilic inflammation but also involve direct restoration of epithelial function.

Aberrant epithelial repair is a key driver of AR and, particularly relevant to severe asthma, contributes to increased susceptibility to viral infections and environmental insults, perpetuating inflammation and recurrent exacerbations. Impaired wound healing has previously been attributed to epithelial alarmins and IL-13^43^. Our findings show that IL-5 also directly impairs epithelial wound repair, providing functional evidence that IL-5 signalling adversely affects epithelial repair capacity. The accompanying shift towards a wound-healing/repair M2 macrophage phenotype further supports a role for IL-5 blockade in promoting repair and remodelling resolution.

The physiological relevance of these observations is supported by our histological data. Consistent with previous reports^8–10^, IL-5 blockade reduced features of AR. We extend these observations by providing a mechanistic link between epithelial reprogramming and structural improvement and, for the first time, demonstrating enhanced epithelial barrier integrity, evidenced by increased expression of E-cadherin and improved cell-cell communication networks.

Our findings support the concept that AR is not solely determined by eosinophilic inflammation. Previous studies have reported persistent remodelling despite reductions in airway eosinophilia^44^, while IL-13 blockade reduces subepithelial fibrosis without altering tissue eosinophil numbers^45^. Our data provide novel mechanistic evidence that anti-IL-5 therapy inducecs epithelial reprogramming that contributes to improvements in AR.

The sample size reflects the challenges of paired bronchoscopic sampling in severe asthma but was sufficient for robust comparative single cell analyses. Eosinophils were not retained within the single cell dataset, preventing direct analysis of eosinophil-epithelial interactions and the *in vitro* epithelial model does not fully reflect the complexity of the asthmatic airway. However, the integration of single-cell transcriptomics, cell-cell communication analyses, functional epithelial assays and paired histological assessment provides complementary evidence supporting a direct role for IL-5 blockade in epithelial repair and AR.

In conclusion, the effects of IL-5 blockade extend beyond eosinophil suppression and include direct modulation of bronchial epithelial biology. By influencing epithelial transcriptional programmes, repair responses and intercellular signalling networks, IL-5 blockade attenuates key features of AR, including impaired epithelial integriy, BM thickening and goblet cell hyperplasia. These findings broaden the current understanding of IL-5 biology, provide evidence that AR in severe asthma is modifiable and highlight the airway epithelium as an important mediator of treatment response in severe asthma.

## Author contributions

Conception and funding acquisition: HR, RTMN, EJS, CB. Methodology: HR, RTMN, EJS, CB, OZ. Sample acquisition and analysis: all authors. All authors discussed the results. Writing: OZ, RTMN, EJS, CB, HR with input from all authors.

## Data availability statement

All single cell data is accessible in GEO and reviewer links are available. The datasets will also be incorporated to the Human Lung Cell Atlas.

## Ethical approval

REC reference 21/EE/0228, IRAS 304497, ERGOII reference 68680.A1.

## Patient consent statement

All patients provided informed written consent

## Clinical trial registration

NCT05144087

## Permission to reproduce material from other sources

N/A.

## Acknowledgment

The authors would like to acknowledge Ms Naomi Banan, lead respiratory research nurse for this study.

