## Supplemental Methods, Table and Figures for "IL-5 blockade restores the bronchial epithelium and attenuates airway remodelling in severe asthma"

### Supplementary Materials and Methods

#### Bronchial biopsy and brushes: collection and preservation

Bronchial brushes and biopsies were collected from the bronchus intermedius and third and fourth airway carinae respectively under direct vision. Bronchial brushings (attached to the brush) were immediately transferred into ice-cold freezing medium (Airway Epithelial Cell Growth Medium (#C-21160, PromoCell, Heidelberg, Germany) and placed in a Coolcell on dry ice and transferred to -70°C overnight prior to cryopreservation in liquid nitrogen while bronchial biopsies were formalin fixed, paraffin embedded (FFPE) and stored prior to further analysis. Do you want to say from the bronchus intermedius and 3rd and 4th airway carinae respectively, to identify where the samples were taken from

#### Brushings processing for single cell

Bronchial brushes were thawed and immediately transferred in room temperature RPMI 1640 Medium, GlutaMAX (Gibco, 11554516) enriched with 6.5 mM DTT (Thermo Scientific, 10699530). An equal volume (10 ml) of StemPro Accutase (Gibco, 11599686) was added and the cell suspension was incubated at room temperature for 5 minutes under gentle swirling. After incubation, 20 ml of serum-free RPMI were added to quench Accutase activity and the cell suspension was filtered through 70 μm cell strainers. The suspension was then centrifuged at 400 RCF for 10 minutes at room temperature. After decanting the supernatant, the resulting pellet was resuspended in 500 μL of PBS with 0.04% BSA (Bio-Techne, 5217/100G). Cells where then tested for viability, presence of clusters and debris and diluted accordingly for single cell Gel Bead-in-Emulsion (GEM) generation following 10X Genomics technologies instructions.

#### Single cell analysis

Single-cell RNA sequencing libraries were generated using 10X Genomics Chromium platform with the Next GEM Single Cell 3’ Gene Expression, v3.1 kit. Libraries were sequenced according to the manufacturer's protocol. Libraries were sequenced by the Genomics and Microarray Core facility of University of Colorado. Raw fastq files were processed using CellRanger (v13.2.0; 10X Genomics) for demultiplexing, alignment to the reference genome, barcode assignment, and generation of gene-cell count matrices. Downstream analyses were performed in RStudio v4.4.1. Quality control, normalisation, dimensionality reduction, clustering, differential gene expression analysis and visualisation were performed using Seurat v5.4.0[1]. Integration of the datasets and differential expression were performed using respectively Harmony v1.2.4 Rcpp_1.1.1[2] and DESeq2 v1.44.0[3]. Plotting and trajectory analysis were performed with the R package SeuratExtend v1.2.4[4]. Cell to cell communication analysis was performed using CellChat v2.1.2[5].

#### Bronchial epithelial cell (BEC) culture

BEAS-2B cells: To generate BEAS-2B cells stably expressing IL-5RA, lentiviral particles were produced by transient transfection of HEK-293T packaging cells. Briefly, $2\times10^6$ HEK-293T cells were seeded on 10 cm dishes and transfected the following day with a three-plasmid system comprising the IL5RA expression vector (Vector

Builder, pLV[Exp]-TagBFP2-EF1A>{6His-IL5RA}, VB250428-1232cjj) the second-generation packaging plasmid p8.91 encoding GAG and POL, and the VSV-G envelope plasmid pMD2.G, using Lipofectamine. Virus-containing supernatant was collected 48 hours post-transfection, filtered through a 0.45 µm membrane, and centrifuged at 400 RCF for 15 minutes. BEAS-2B target cells were pre-incubated with RPMI with GlutaMAX (ThermoFisher Scientific, 61870036) supplement and further supplemented with polybrene (final concentration 8 µg/ml) for 30 minutes prior to transduction. Cells were transduced with clarified viral supernatant by centrifugation at 1200 RCF for 90 minutes at 37°C. Transduction was repeated the following day.

BCi.NS1.1 cells: BCi-NS1.1[6] (BCi herein) cells were kindly gifted by Prof Crystal (Weill Cornell Medical College) and continuously cultured as adherent monolayers in collagen I (30 µg/ml) pre-coated culture flasks with basal media (PneumaCult™-Ex Plus Medium, Stemcell Technologies, 05040).

Primary differentiated BECs: These were obtained commercially from Epithelix. Air-Liquid Interface (ALI) cultures from healthy donors were sustained for 24 hours in MucilAir- culture medium (Epithelix, EP01MD) and subsequently apically and basally exposed to IL-5 (10 ng/ml, Biotechne, 205-IL-005/CF) for 24 hours. Transwells were then washed twice with PBS and cells were lysed with RIPA buffer (50 mM Tris-HCl, pH 7.4; 150 mM NaCl; 1.0% NP-40; 0.5% Sodium deoxycholate; 0.1% SDS; 1 mM EDTA) enriched with proteinase inhibitors (Cell Signaling Technology, 5871S).

**Scratch wound assays**
For differentiated ALI cultures, BCi cells were seeded ($0.15x10^6$/well) onto collagen I (30 µg/ml) pre-coated 0.4 µm porous membranes and submerged for 24h in basal media prior to culture at the ALI for 28 days in ALI media (PneumaCult™-ALI medium, Stemcell Technologies, 05001). Seven days post-ALI culture, IL-5 (0.5 or 10ng/ml, Biotechne, 205-IL-005/CF) was added for the remaining 21 days. After which, ALI cultures were scratched with a 200 µl pipette tip and repair of the wound in the presence or absence of IL-5 (0.5-10 ng/ml) was visualised in 3 views per membrane over 24h at 3h intervals using time-lapse light microscopy at 37°C in 5%$CO_2$ (Leica). Percent of wound closure was calculated from t=0.

**Western blotting**
Protein sample was mixed with loading buffer to 1x, added of β-mercaptoethanol 5% and heated at 90 °C for 5 minutes. The sample was then resolved in NuPAGE™ Bis-Tris Mini Protein Gels 4-12% (Invitrogen, NP0321) and blotted on nitrocellulose membrane via mini blot module (Invitrogen 10572913). The membrane was then blocked in TBS, 0.05 % Tween and 5% ECL blocking agent (Fisher Scientific, RPN2125). Primary antibody incubation using anti human IL5Rα (Bio-Techne, AF-253-NA) was performed overnight at 4 °C in blocking buffer. The membrane was then washed three times with TBS+0.05% Tween and incubated with HRP-conjugated secondary antibody (Abcam, ab97110) for 1 hour at room temperature. The membrane was washed again before being treated with SuperSignal West Pico PLUS Chemiluminescent Substrate (Thermo Scientific, 34578) for imaging.

**Histology and Immunohistochemistry**

Sections (10 µm) of endobronchial biopsy sections (fixed in 4% paraformaldehyde and paraffin embedded) were processed and stained as previously described (XX). Briefly, the biopsies sections were de-waxed, rehydrated and incubated in 0.5% hydrogen peroxide in methanol for 10 min to block endogenous peroxidase activity. Following antigen retrieval (0.05% pronase for 10 min at room temperature, 0.01M citrate buffer or 1mM EDTA buffer for 25 min in microwave depending on primary antibody used), sections were then blocked with avidin/biotin blocking solution (Vector SP-2001) followed by blocking medium (10% FBS+DMEM) before incubated overnight at 4°C with a primary antibody against ECP (1:500, Diagnostics development mAb593), collagen I (1:100, Abcam ab6038), or e-cadherin (1:300, Abcam Ab40772). After washing, bound antibody was detected using a biotinylated secondary conjugated antibody (either goat anti-rabbit IgG (1:800, Vector BA-1000) or goat anti-mouse IgG (1:800, Vector BA-9200) depending on primary antibody) followed by avidin-biotinylated horseradish peroxidase (Vector PK-6100) and visualisation using DAB. Sections were counterstained with Mayer's Haematoxylin. In addition, adjacent serial sections were stained using Movat's Pentachrome stain. Images were acquired using a Zeiss microscope and software (Zeiss KS400.30). For quantifying eosinophils, cell numbers in the submucosa were divided by the submucosal area ($mm^2$) to give cells/$mm^2$. For quantifying remodelling and epithelial markers, %positive staining as a % of total biopsy area ($mm^2$) was calculated while for epithelial barrier markers only endobronchial biopsies with intact epithelium were used. Analyses were performed by an observer blinded to the treatment stage of the participants.

### Statistical Analysis

Statistical analysis of clinical parameters, immunohistochemistry and scratch wound assays was performed in GraphPad Prism v10.3.1 (GraphPad Software Inc, San Diego, CA). Normality of distribution was assessed using the Shapiro-Wilk normality test; paired Student's t test or Mann-Whitney U test were used to assess differences between pre- and post-anti-IL-5 treatment for clinical parameters and immunohistochemistry analyses in parametric or non-parametric data respectively. Differences between scratch wound repair in differentiated BCi cells in the presence or absence of IL-5 was assessed using a 2-way ANOVA with Tukey correction for multiple comparisons. Results were considered significant if $P \leq 0.05$ where *$P \leq 0.05$, **$P \leq 0.01$and ***$P \leq 0.001$.

**Supplementary Table S1.**

| **Clinical Parameter (median, IQR)** | **Baseline, pre-anti-IL-5 treatment** | **6 months post anti-IL-5 treatment** | **Median of differences** | **p-value** |
|---|---|---|---|---|
| **Exacerbations (annualised exacerbation rate)** | 5 (4,6) | 0 (0,0) | -5 | 0.0001 |
| **ACQ-6 score** | 2.65 (2.10,3.08) | 0.80 (0.30,2.00) | -1.40 | 0.001 |
| **Blood eosinophil count (x$10^9$/L)** | 0.45 (0.33,0.70) | 0.10 (0.0,0.10) | -0.40 | <0.001 |
| **FeNO (ppb)** | 47 (33,79) | 56 (33,97) | +13 | 0.25 |
| **Pre-BD FEV1 (L)** | 1.89 (1.62,2.45) | 2.10 (1.70,2.77) | +0.11 | 0.08 |
| **Pre-BD FEV1 %predicted** | 63.50 (58.75,85.00) | 75.00 (61.75,86.50) | +4 | 0.02 |
| **Pre-BD FVC (L)** | 3.13 (2.92,3.80) | 3.42 (3.74,3.79) | 0 | 0.94 |
| **FEV1/FVC (%)** | 60.00 (56.50,68.75) | 64.50 (58.00,76.00) | +3.5 | 0.02 |
| **Post-BD FEV1 (L)** | 2.15 (1.85,2.68) | 2.28 (2.0,2.85) | 0.07 | 0.13 |
| **BAL eosinophils (%)** | 3.67 (0.44,7.38) | 0.63 (0.19,2.15) | -1.67 | 0.04 |

Table S1: Clinical characteristics of participants at baseline and after 6-months of anti-IL-5 treatment

ACQ: asthma control questionnaire; BAL: bronchoalveolar lavage; BD: bronchodilator FeNO: fraction exhaled nitric oxide; FEV1: forced expiratory volume in 1 second, FVC: forced vital capacity

### Supplementary Figures

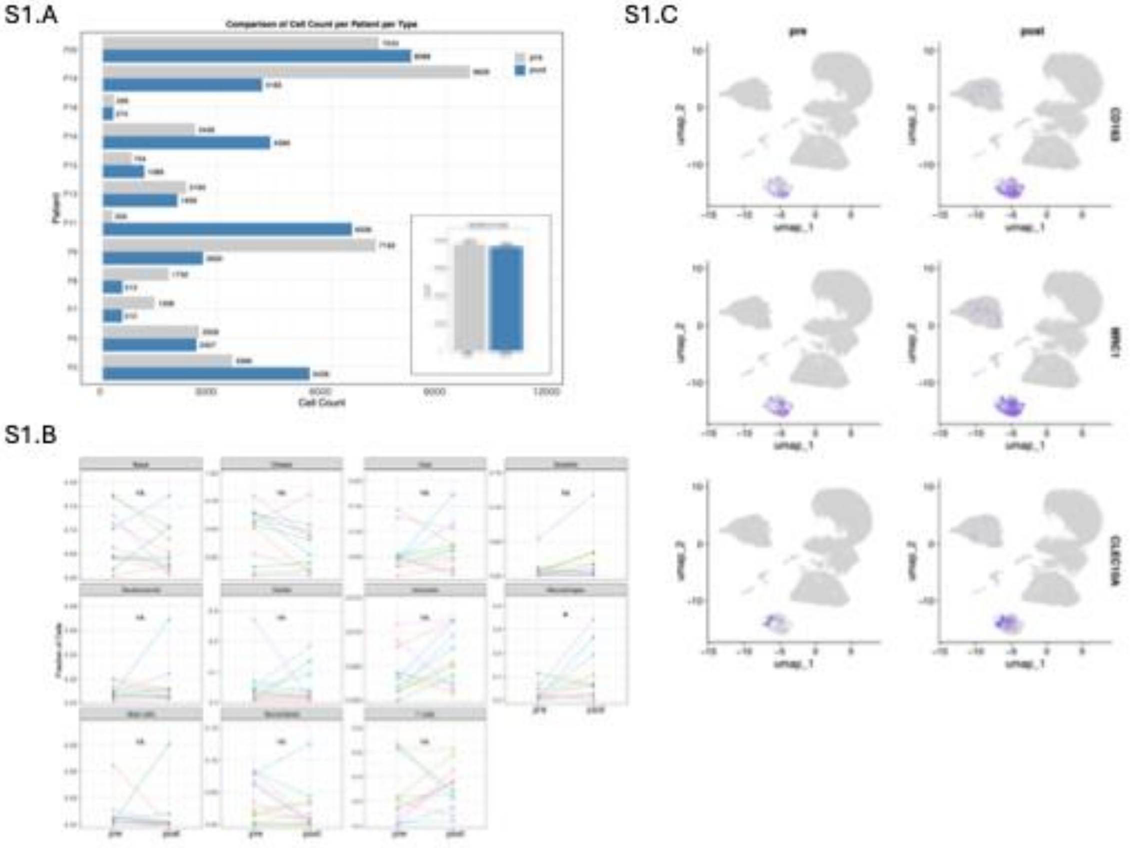


**Figure S1. S1A**. Bar plot showing the number of cells analysed per patient pre- and post-treatment. Internal bar plot shows the total number of cells analysed pre- and post-treatment. **S1B**. Plots showing inter-individual changes in the sample composition pre and post treatment for each cell type found. **S1C**. UMAP plots highlighting the cells expressing marker genes (*CD163*, *MRC1/CD206* and *CLEC10A/CD301*) of M2-type macrophages, enriched post treatment.

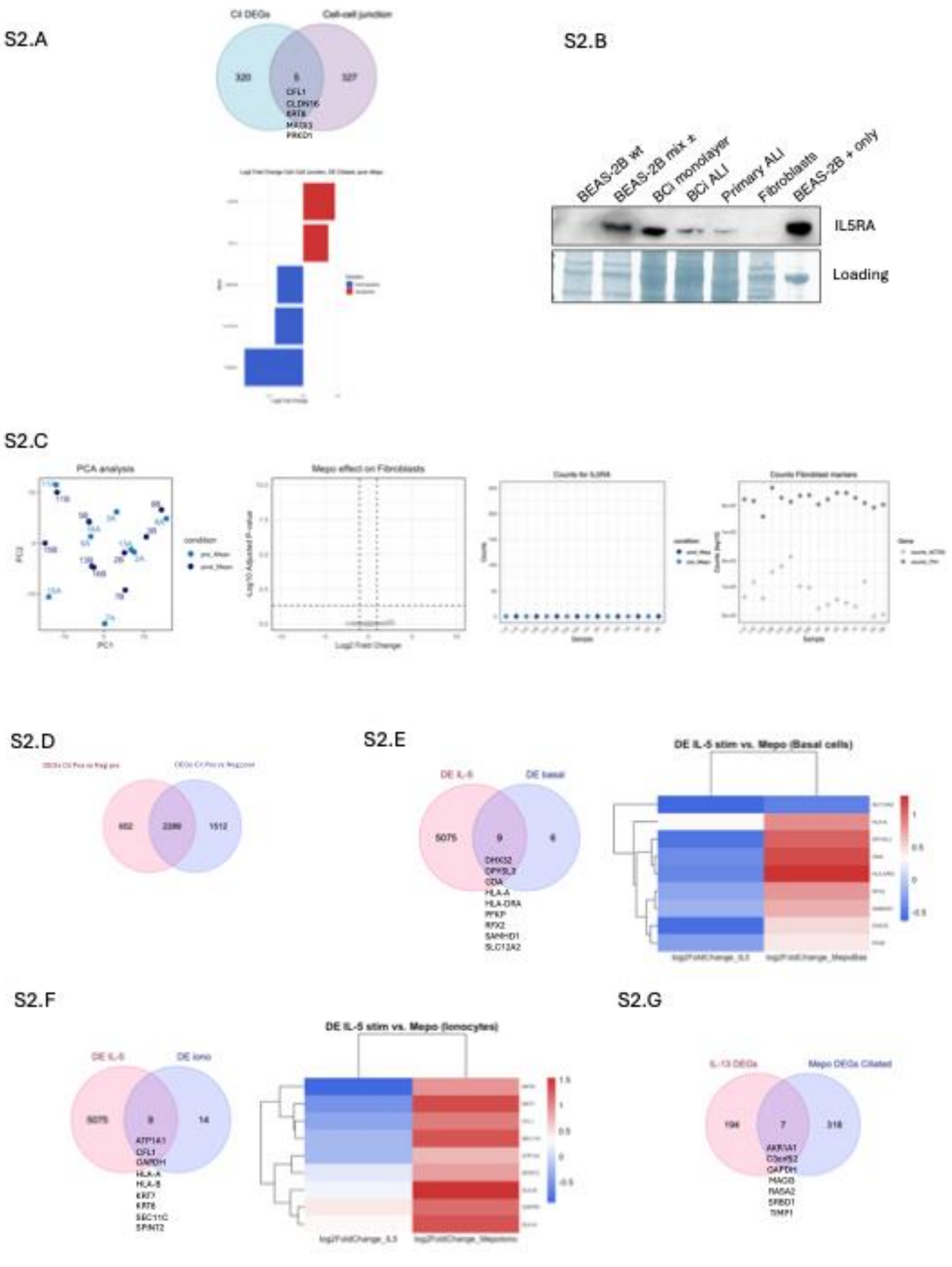


**Figure S2. S2A**: Top: Venn diagram showing the overlap between DEGs in ciliated cells post-IL-5 blockade and genes within the biological term “Cell-cell junction”. Bottom: bar plot listing specific genes in the biological term and showing the direction (blue, downregulated and red, upregulated) and magnitude (as $\log_2$ Fold change) of the changes. **S2B:** Western blot for IL-5Rα in control wild type BEAS-2B cells (BEAS-2B wild type (wt), lane 1), mix of BEAS-2B wild type and transgenic for *IL5RA* (Mix BEAS-2B ±, lane 2), BCi cell monolayers (BCi monolayer, lane 3), BCi cells differentiated at the ALI (BCi ALI, lane 4) primary BEC ALI cultures (Primary ALI, lane 5), SEA fibroblasts (Fibroblasts, lane 6) and BEAS-2B cells overexpressing *IL5RA* (BEAS-2B + only, lane 7). Ponceau staining shown as a loading control. **S2C**: From left to right: PCA analysis of bulk RNA-seq data of SEA fibroblasts outgrown from bronchial biopsies pre- and post-IL-5 blockade in vivo. Volcano plot showing the lack of significant (adjusted p-value, padj $< 0.05$) DEGs in SEA fibroblasts pre- and post-IL-5 blockade. Dot plot showing the number of counts from bulk RNA-seq data relative to *IL5RA* in SEA fibroblasts. Dot plot showing the number of counts (expressed as $\log_{10}$ of the value) from bulk RNA-seq data relative to fibroblast marker genes (*ACTA2* and *FN1*) in SEA fibroblasts analysed. **S2D:** Venn diagram showing the overlap (2289) between all DEGs obtained from the comparison between ciliated cells positive for the expression of *IL5RA* pre-treatment (2941) and post-treatment (3801). **S2E:** Left. Venn diagram showing the overlap between the DEGs induced by IL-5 treatment of ALI culture[1] and DEGs found in basal cells post IL-5 blockade. The genes overlapping are listed. Right. Heatmap comparing common DEGs obtained from exposure of ALI cultures to IL-5[1] and DEGs found in basal cells post IL-5 blockade. **S2F** Left. Venn diagram showing the overlap between the DEGs induced by IL-5 treatment of ALI cultures[1] and DEGs found in ionocytes post IL-5 blockade. The genes overlapping are listed. Right. Heatmap comparing common DEGs obtained from exposure of ALI cultures to IL-5[1] and DEGs found in ionocytes post IL-5 blockade. **S2G**: The Venn diagram shows the common genes differentially expressed post IL-13 stimulation and IL-5 blockade in ciliated cells.

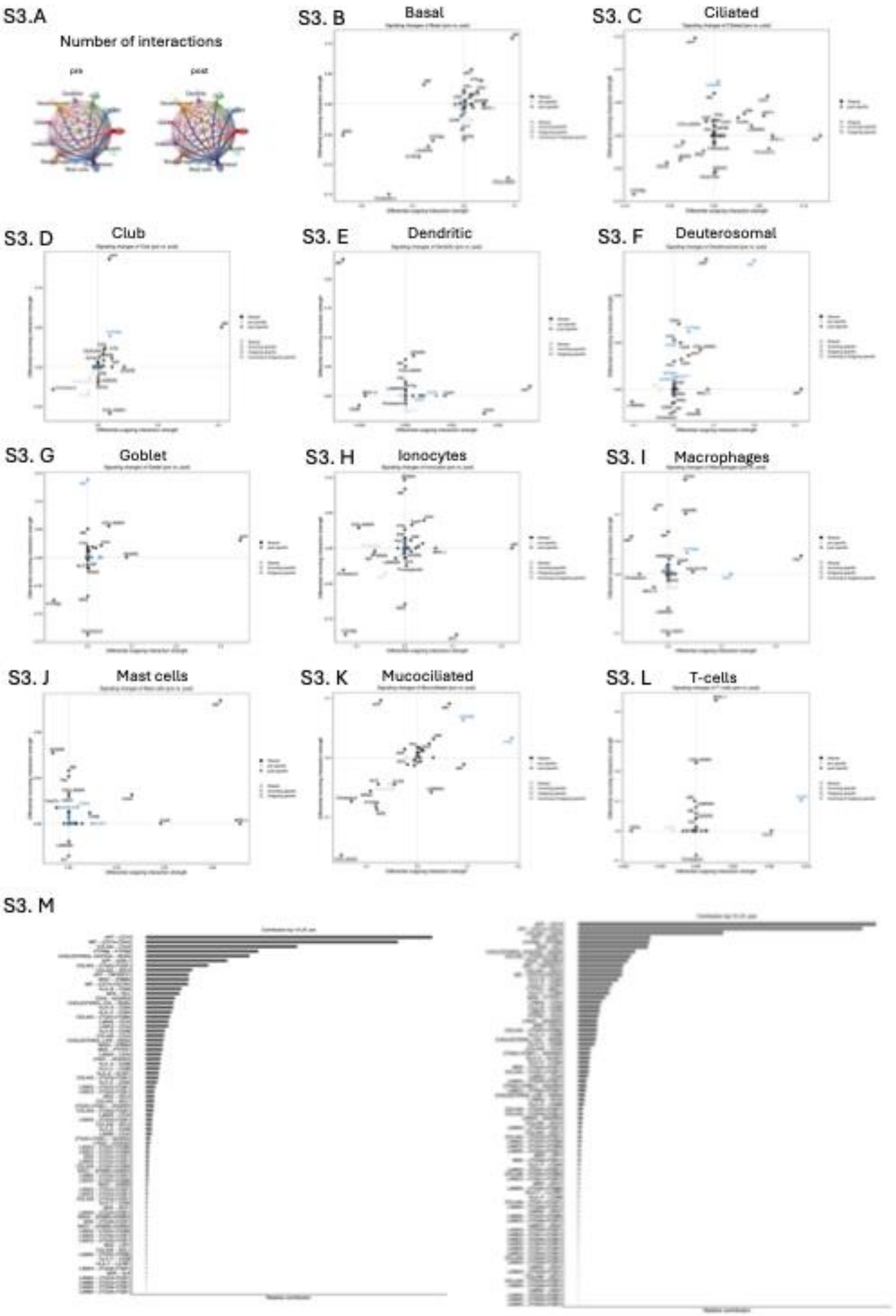


**Figure S3.** Circle plot showing the network of cell-to-cell interactions pre and post IL-5 blockade. **B to L.** Scatter plots showing the pathways involved in signalling changes associated with anti-IL-5 treatment for basal cells (**B**), ciliated cells (**C**), club cells (**D**), dendritic cells (**E**), deuterosomal cells (**F**), goblet cells (**G**), ionocytes (**H**), macrophages (**I**), mast cells (**J**), mucociliated cells (**K**) and T cells (**L**). **M.** Bar plots showing the contribution of the top 10 active pathways pre (left) or post (right) treatment. **N**. Bar plot showing the main pathways active and the proportion of their activation (expressed as information flow) between pre and post treatment.

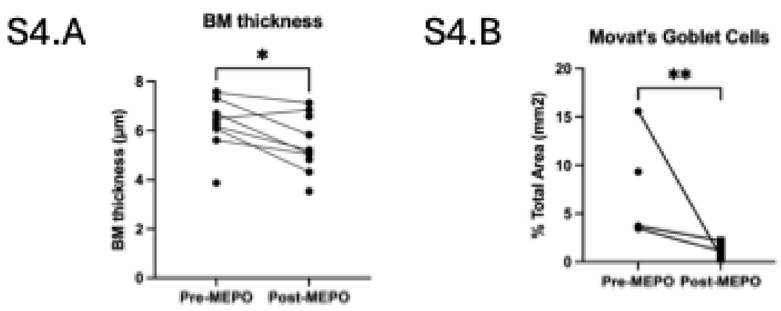


**Figure S4.A**. Dotplot showing the quantification of basal membrane thickness. **S4.B**. Dotplot showing the count of goblet cells post Movat's staining from bronchial biopsy slices pre- and post-IL-5 blockade.
